# The Brain, the Bugs, and the Binge: A Protocol Intervention Examining the Interplay Between Binge drinking, Gut Microbiota, and Brain Functioning

**DOI:** 10.1101/2024.12.12.24318771

**Authors:** Diogo Prata-Martins, Clarisse Nobre, Natália Almeida-Antunes, Pedro Azevedo, Sónia S. Sousa, Alberto Crego, John F. Cryan, Adriana Sampaio, Carina Carbia, Eduardo López-Caneda

## Abstract

**Background:** Adolescence and youth are periods of significant maturational changes which seem to involve greater susceptibility to disruptive events in the brain such as binge drinking (BD). This pattern -characterized by repeated alcohol intoxications- is of special concern, as it has been associated with significant alterations in the young brain. Recent research indicates that alcohol may also induce changes in gut microbiota composition and that these disturbances can lead to impairments in the brain and behaviour. Additionally, there is evidence suggesting that microbiota-targeted interventions (psychobiotics) may have a beneficial impact on mitigating alcohol-induced damage in chronic alcoholics, while also potentially influencing cognitive/brain functioning. However, this triadic relationship between BD, gut microbiota, and brain structure/functioning as well as the therapeutic potential of gut microbiota-targeted interventions in youths with a BD pattern remains largely unexplored.

**Method:** This double-blind, parallel, randomized controlled study (registered in the clinical trials database; NCT05946083) aims to evaluate whether a BD pattern may disrupt gut microbiota balance in young college students. Furthermore, it seeks to determine whether alcohol-induced alterations in the microbial composition and functions are associated with immune, cognitive, neurostructural, and neurofunctional impairments. For this purpose, 82 college students (36 non/low drinkers and 46 binge drinkers [BDs]), matched for age and gender, will be recruited. During the pre-intervention phase, all participants will undergo comprehensive assessments, including microbiota and inflammatory profiling, neuropsychological testing, and evaluations of brain structure and function using magnetic resonance imaging. Subsequently, only BDs will proceed to the intervention phase, which involves a 6-week regimen of either a prebiotic (inulin) or placebo (maltodextrin). Following the intervention, the baseline variables will be re-evaluated and, over the subsequent 3 months, levels of craving and alcohol consumption will be monitored.

**Discussion:** This will, to our knowledge, be the first study to assess the potential relationship between gut microbiota and neurocognitive functioning in young BDs. The administration of psychobiotics is expected to promote the restoration of gut microbiota, thereby decreasing pro-inflammatory responses and mitigating alcohol-induced brain disruptions. This reduction in the detrimental effects of alcohol is anticipated to lead to cognitive enhancements and neurofunctional changes. Collectively, these findings might have major implications for understanding and treating alcohol misuse, potentially leading to new therapeutic strategies targeting the gut-brain axis to alleviate some of the detrimental effects associated with excessive alcohol use.

**Clinical Trial registration:** ClinicalTrials.gov, ID: NCT05946083.

**Strengths and Limitations:** The present protocol has several notable strengths such as:

- <u>Novel insights into the influence of alcohol on the microbiota-gut-brain axis</u>. This study is the first to date to examine the interaction between binge drinking, gut microbiota, and brain function.
- <u>Breakthrough therapeutic impact</u>. This is a double-blind, parallel, randomized controlled trial focuses on a relevant population group -young college students- who are particularly vulnerable to harmful alcohol consumption patterns. By exploring prebiotic interventions, the study introduces innovative strategies to mitigate the neurotoxic effects of alcohol, opening new ways for therapeutic approaches in mental health and preventive medicine.
- <u>Multidisciplinary approach</u>. By integrating analyses of gut microbiota, inflammatory profile, and neuropsychological and neurostructural assessments, this study offers a comprehensive investigation of the impact of binge drinking on the microbiota-gut-brain axis.

However, there are limitations to consider, including:

- <u>Potential for low adherence</u>: Young binge drinkers may have low adherence to a six-week intervention, which could compromise data validity.
- <u>Uncontrolled external factors</u>: Factors such as diet, exercise, sleep, and stress, which can affect both microbiota and brain function, are not fully controlled. This could hinder the interpretation of the results and mask the effects of the intervention.

## 1. INTRODUCTION

A prevalent form of alcohol misuse among young people that has garnered considerable attention in the last two decades is binge drinking (BD), characterized by episodes of excessive alcohol consumption followed by periods of low intake or abstinence (López-Caneda, Cadaveira, et al., 2019; Maurage et al., 2020). According to the National Institute on Alcohol Abuse and Alcoholism (NIAAA), BD is defined as a pattern of alcohol use that results in a blood alcohol concentration (BAC) level of 0.08 g/dL, typically occurring after consuming a minimum of four (for women) or five (for men) alcoholic beverages within a span of two hours, at least once a month (NIAAA, 2020).

Concern over the rising prevalence of BD among young people has emerged as a critical public health issue, particularly due to the heightened vulnerability of this age group to the neurotoxic effects of alcohol (Jones et al., 2018). In most Western countries, including Portugal, this high consumption pattern is a regular practice in approximately one-third of youths aged 15-24 years (Group et al., 2015). The implications of this behaviour extend beyond immediate physical and quality-of-life issues, encompassing neurocognitive consequences (Almeida-Antunes et al., 2021; Dormal et al., 2018; White & Hingson, 2013). Research focusing on the long-term impacts on young binge drinkers (BDs) has identified alterations in both brain structure and function, highlighting the severity and complexity of the risks associated with excessive alcohol use in this stage of development (Lees et al., 2019; Pérez-García et al., 2022).

Accumulating evidence indicates that alcohol misuse induces inflammation not only by acting directly on the brain –e.g., through proinflammatory cytokines-but also through systemic interactions, for example via the gut microbiota (Carbia et al., 2021; Jadhav et al., 2018). The gut microbiota, comprises over 100 trillion microorganisms primarily residing in the lower gut, existing in a symbiotic relationship with the host (Kho & Lal, 2018). It plays a crucial role in nutrient metabolism, fermenting non-digestible substrates such as prebiotics and dietary fiber to produce microbial metabolites, such as short-chain fatty acids (SCFAs). Additionally, it regulates immune function and contributes to the production of neurotransmitters, including serotonin, gamma-aminobutyric acid (GABA), and dopamine, all of which have significant implications for overall homeostasis regulation (Clarke et al., 2014; Hillemacher et al., 2018; Temko et al., 2017).

Recent research has underscored the pivotal role of gut microbiota in mental health conditions, including alcohol use disorders (Carbia et al., 2021; Stärkel et al., 2016). Accordingly, chronic alcohol use has been linked to changes in gut barrier function, resulting in disruptions in intestinal permeability (leaky gut) which, consequently, can lead to the leakage of bacteria, toxins, and other bacterial products, such as lipopolysaccharides (LPS), through the gut and causing inflammation in both the gut and liver. This peripheral inflammation may contribute to brain inflammation (de Timary et al., 2017). Elevated levels of pro-inflammatory cytokines (e.g., the interleukins IL-1β, IL-6, and IL-8, and the Tumour Necrosis Factor α [TNF-α]) or activated immune cells in the bloodstream can cross the blood-brain barrier, inducing inflammation in microglia or astrocytes(de Timary et al., 2017). Certain bacterial products from gut metabolism, such as tryptophan metabolites, have also been shown to significantly influence brain inflammation (Rothhammer et al., 2016). In addition to the production of circulating inflammatory cytokines, alcohol consumption has been associated with metabolic alterations, including a decrease in the production of immunoglobulin A (IgA) and SCFAs, both crucial in the regulation of microglia in the central nervous system(Zheng et al., 2020). Altogether, these cumulative effects highlight the potential for alcohol abuse to disrupt the delicate balance between gut microbiota, immune responses, and brain health.

While inconsistencies exist, research on alcohol-related microbiome alterations suggests the presence of specific microbial taxonomic signatures linked to dysfunctions associated with excessive alcohol use. Accordingly, preclinical studies showed decreased gut microbial α- and β-diversity —(i.e., diversity within and between microbial communities, respectively)- in animals exposed to alcohol intake (Hofford & Kiraly, 2024). In addition, some studies in animal models have reported differential relative abundance at the phylum level, exhibiting reductions in Bacteroidetes and increase in Firmicutes, i.e., an elevated Firmicutes/Bacteroidetes ratio (Chivero et al., 2022; X. Wang et al., 2023). On the other hand, research conducted on patients with alcohol use disorder (AUD) has reported reductions in α-diversity, as well as a depletion of beneficial bacteria from the Bacteroidetes and Firmicutes phyla (including the Ruminococcaceae and Lachnospiraceae families), alongside an increase in bacteria linked to pro-inflammatory activity, such as Proteobacteria (Addolorato et al., 2020; Du et al., 2022; Leclercq et al., 2014; Mutlu et al., 2012). Although data is limited, preclinical research investigating the link between gut microbiota and binge drinking has also indicated microbial dysbiosis induced by excessive alcohol intake in rats, which persisted into adulthood (Vetreno et al., 2021). Similarly, a recent study involving young BDs identified alterations in their gut microbiota, which were in turn associated with alcohol craving and emotional and cognitive difficulties (Carbia et al., 2023).

Altogether, considering the relationship between alcohol use disorder (AUD) and gut microbiota alterations as well as the recent research showing gut microbiota changes in youths with heavy alcohol drinking (Carbia et al., 2023; Segovia-Rodríguez et al., 2022), gut microbiota-targeted interventions may constitute a promising approach for developing new therapeutic interventions, not only to minimize alcohol-induced damage (Hillemacher et al., 2018; Leclercq et al., 2020) but also to modify alcohol use patterns (Amadieu et al., 2022; Bajaj et al., 2021; Ezquer et al., 2022). In this context, prebiotics, probiotics, synbiotics, fecal microbiota transplants, and specific dietary interventions emerge as promising therapeutic options, increasingly recognized for their documented benefits on mental health (Berding & Cryan, 2022; Dinan et al., 2013; Kazlausky Esquivel, 2021; Kim & Mills, 2024). Prebiotics, which are non-digestible substrates that selectively stimulate the growth and activity of beneficial bacteria in the gut (Manning & Gibson, 2004), have been shown to significantly influence the microbiota-gut-brain axis. This includes improvements in cognitive functions (Berding et al., 2021) and mental health (Cryan et al., 2020), beneficial changes in gut microbiome (Enam & Mansell, 2019; S. Wang et al., 2020), and derived positive metabolic outcomes such as increase in SCFAs or circulating brain-derived neurotrophic factor (Amadieu et al., 2022; O’Riordan et al., 2022; Savignac et al., 2013)

Thus, considering that any gut microbiota disruption may exacerbate neuroinflammatory conditions and contribute to the progression of neurological disorders, it’s important to maintain gut health to preserve overall brain function and mitigate the impacts of alcohol misuse on neurological health. Furthermore, as the effects of BD on gut microbiota is still a relatively unexplored research field, gut microbiota-targeted interventions may eventually improve the composition and functioning of the gut microbial community and thereby reduce the neurotoxic effects associated with excessive alcohol consumption. Accordingly, the present protocol aims to define the procedures for exploring the gut microbiota and its potential relationship with brain structure and function in young BDs. The goal is to develop and implement a randomized, placebo-controlled trial that evaluates gut microbiota targeted at reducing alcohol misuse and associated harm.

## 2. AIMS AND HYPOTHESES

The main objectives of this multidisciplinary study are: O1) (Binge & Bugs) to evaluate whether a BD pattern of alcohol consumption may induce gut microbiota alterations in young college students; O2) (Bugs & Brain) to determine whether the alcohol-induced altered microbial profile is associated with immune, cognitive, neurostructural and neurofunctional impairments in young BDs; O3) (Bugs, Brain & Binge) to examine whether administration of prebiotics (inulin) -through a randomized control trial (RCT)- in young BDs improves the composition of the gut microbiota and, consequently, reduces the alcohol-induced cognitive/brain anomalies. Therefore, this experimental procedure aims to answer three main research questions and test specific hypotheses:

### i. Can BD induce gut dysbiosis in young college students?

It is hypothesized that, compared to age-matched controls (i.e., non/low drinkers), BDs will exhibit disruptions in the gut microbiota composition. Possibly replicating the findings of Carbia at the baseline level and expanding on these baseline changes and their relation to brain function (Carbia et al., 2023), BDs will essentially (though not exclusively) display a reduction of gut diversity and/or disruptions in bacterial richness (e.g., reduced levels of Bacteroidetes and Firmicutes phyla, and increased abundance of Proteobacteria).

### ii. Is alcohol-induced gut dysbiosis associated with increased immune response and/or neurocognitive and brain structure/functioning impairments in young BDs?

It is expected that BDs show augmented plasma levels of pro-inflammatory cytokines (e.g., IL-1β, IL-6, TNF-α) (Claesson et al., 2012; Orio et al., 2018; Pascual et al., 2017). Additionally, and based on previous literature, microbiota profile will tentatively be associated with neuropsychological performance (Carbia et al., 2023), brain morphometry measures (e.g., amygdala volume; (Maki et al., 2024) as well as brain functioning linked to emotional processing and executive functioning (Ahluwalia et al., 2014; Bagga et al., 2018; Tillisch et al., 2017)).

### iii. Will the administration of prebiotics in young BDs improve the composition of the gut microbiota and, consequently, reduce alcohol-induced cognitive/brain anomalies?

The administration of prebiotics is anticipated to promote the restoration of the gut microbiota (primary outcome), e.g., as indicated by increased α-diversity and positive bacterial richness compared to pre-intervention assessment. This restoration is expected to mitigate alcohol-induced brain damage in young BDs, with anticipated improvements across various secondary outcome domains. These domains include immunological (reduction of pro-inflammatory markers), cognitive (enhancement in neuropsychological performance), and neurofunctional levels (e.g., diminished neural reactivity to alcohol cues).

## 3. MATERIALS AND METHODS

### 3.1. Study Design and Setting

The present protocol was registered in the clinical trials database of the National Institutes of Health (NIH) (identifier: NCT05946083). The participant recruitment started in March 2023 and is expected to be completed by July 2025. The SPIRIT checklist is available in the Supplementary Material. The study comprises several stages, each aimed at ensuring the quality and reliability of the results (see Figure 1, Image files document).

**Figure 1.**
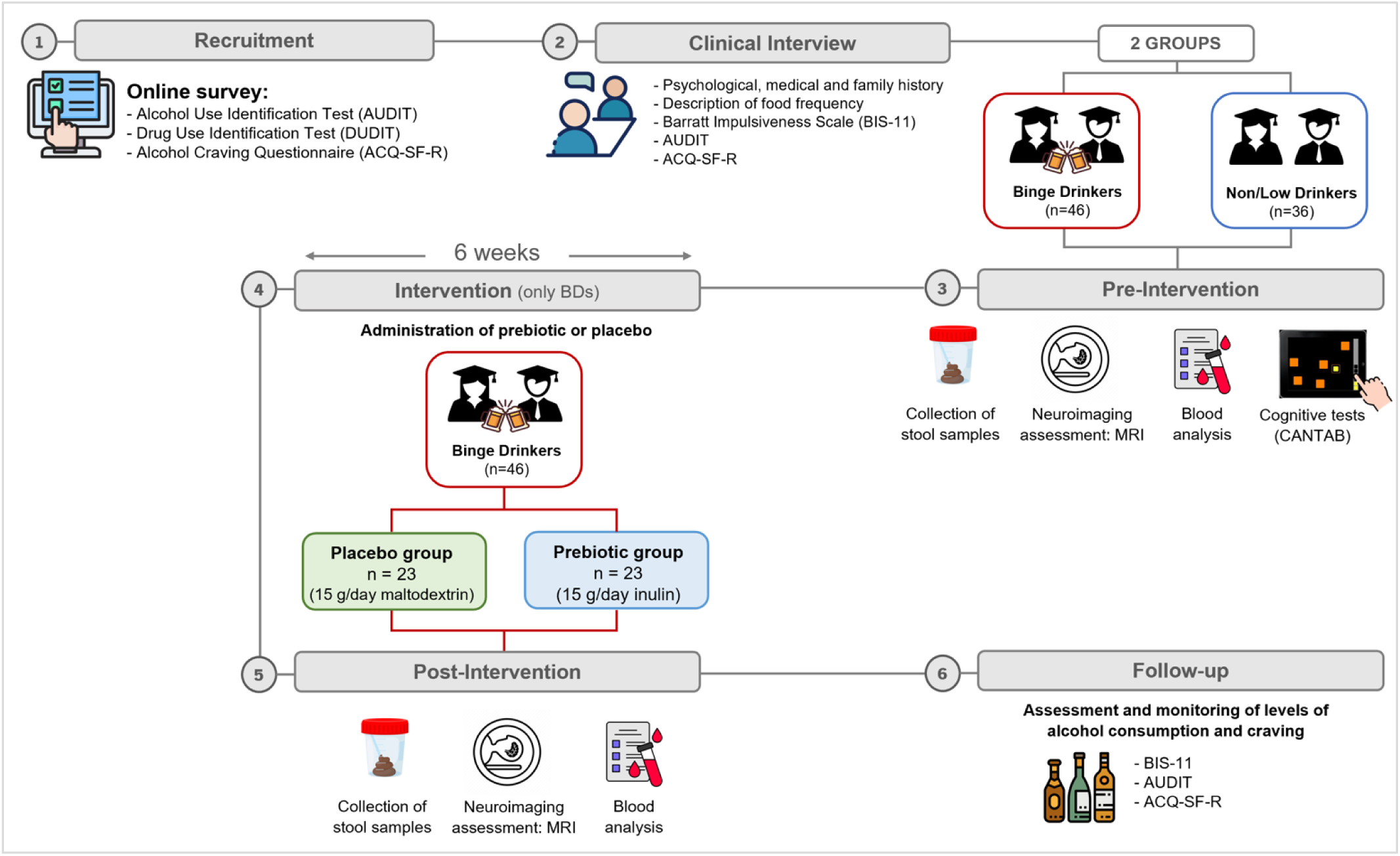
Study outline. Eighty-two participants will be recruited: 36 non/low drinkers and 46 binge drinkers (BDs), matched for age and gender. Recruitment will be carried out through an online survey. After sample selection, participants will undergo a clinical interview covering psychological, medical, personal, and family history. According to their drinking patterns, participants will be randomly assigned to two groups: the control group (non/low drinkers) and the intervention group (BDs). During the pre-intervention phase, all participants will be assessed for the variables of interest through neuropsychological tests, collection of stool and blood samples, and magnetic resonance imaging recordings. Subsequently, only the BDs will proceed to the intervention phase, which involves taking either a prebiotic or placebo for 6 weeks. After this period, the variables assessed in the pre-intervention phase will be re-evaluated and, over the following 3 months, the levels of alcohol consumption and craving will be monitored.

#### 3.1.1. Recruitment

The recruitment will be carried out through an online survey broadcasted using the institutional email, which will include items related to alcohol consumption (e.g., frequency, intensity, and alcohol use). A detailed alcohol/drug use will be obtained through several questionnaires: the Alcohol Use Disorders Identification Test (AUDIT; (Babor et al., 2001)), the Drug Use Disorders Identification Test-Extended (DUDIT-E; (Berman et al., 2007)), and the Alcohol Craving Questionnaire - Short Form Revised (ACQ-SF-R; (Rodrigues et al., 2021)). If this approach does not yield the desired number of participants, we will enhance our efforts by conducting face-to-face visits to classrooms, to promote the study directly to young college students and encourage their participation.

#### 3.1.2. Clinical Interview

A detailed clinical interview will be conducted at the School of Psychology of the University of Minho (EPsi, UM) to understand the clinical history and eligibility of the participants. Questions regarding psychological, personal, family, and clinical history will be addressed, including questions about alcohol and drug use history, alongside specific questionnaires related to substance use (i.e., AUDIT, ACQ-SF-R), as well as those related to some psychological traits (e.g., Symptom Checklist-90-Revised questionnaire [SCL-90-R;(Derogatis & Savitz, 1999), Barratt Impulsivity Scale (Patton et al., 1995). After this stage, and according to their drinking patterns, participants will be assigned to two groups: the control group (non/low drinkers) and the intervention group (BDs).

#### 3.1.3. Pre-intervention

The pre-intervention phase involves a comprehensive neuropsychological assessment of the participants, including tasks from the Cambridge Neuropsychological Test Automated Battery (CANTAB) (Cambridge Cognition, UK), aimed to evaluate the emotional and cognitive abilities (see Table 1). The tasks, administered via an iPad, will be conducted at the EPsi (UM), on the same day as the clinical interview, lasting 60 minutes.

**Table 1.** Summary of the cognitive tasks from the Cambridge Neuropsychological Tests Automated Battery (CANTAB).

| Task | Cognitive dimensions | Assessed cognitive functions | Duration |
| --- | --- | --- | --- |
| *Delayed Matching to Sample (DMS)* | Memory | Visual matching ability;<br>Short-term visual recognition memory. | 7 minutes |
| *Emotion Recognition Task (ERT)* | Emotion and Social Cognition | Emotional recognition: ability to identify six basic emotions (sadness, happiness, fear, anger, disgust, and surprise) in facial expressions. | 6 minutes |
| *Cambridge Gambling Task (CGT)* | Executive Function | Decision-making;<br>Risk behaviour outside a learning context. | 12 minutes |
| *Intra-Extra Dimensional Set Shift (IED)* | Executive Function | Cognitive flexibility. | 7 minutes |
| *Spatial Working Memory (SWM)* | Executive Function | Working memory strategies and capacity. | 6 minutes |
| *Stop Signal Task (SST)* | Executive Function | Response inhibition/impulse control | 14 minutes |

In order to establish a robust database for comparative analysis throughout the study, stool and blood samples will be collected for gut microbiota diversity/composition analysis of gut microbiota, and quantification of peripheral levels of interleukins IL-1, IL-6, and IL-8, and TNF-α, respectively, as well as magnetic resonance imaging [MRI] recordings to obtain a neurostructural and neurofunctional evaluation of each participant (see section 4. Assessment Protocol, topics 4.2. Gut Microbiota and 4.3. Neuroimaging). These samples will be collected from each participant in a time interval of less than 72 hours.

#### 3.1.4. Intervention

The BD group will be randomly assigned to one of two intervention subgroups, namely prebiotic or placebo intervention. Inulin (Frutafit^®^ IQ from Sensus B.V.), a well-established prebiotic fiber (Hughes et al., 2022; Shoaib et al., 2016), will be administered as the prebiotic. Maltodextrin, which has a taste and appearance similar to inulin but is not expected to impact the regulation of the intestinal microbiome (Almutairi et al., 2022), will be used as the placebo. Depending on their assigned group, each participant will receive one of two types of fiber over six weeks, with a daily dose of 15g (divided into three doses per day) (Dewulf et al., 2013; Leyrolle et al., 2021).

To uphold scientific standards and prevent bias in results, the study will be conducted under a double-blind design regarding participant group assignments. During each intervention week, dietary habits will be recorded using a semiquantitative food frequency questionnaire (Lopes, 2000; Lopes et al., 2007), documenting all food and beverages consumed over three days: two weekdays and one weekend day.

#### 3.1.5. Post-intervention

The post-intervention phase will initiate with a new collection of stool and blood samples, along with MRI recordings, aimed at identifying potential changes in the microbiota-gut-brain axis resulting from the intervention protocol.

#### 3.1.6. Follow-up

The follow-up phase will extend over three months post-intervention, focusing on assessing and monitoring levels of alcohol use and craving. During this period, BDs will complete specific questionnaires related to alcohol use (e.g., AUDIT, ACQ-SF-R), as well as those related to psychological symptoms (e.g., SCL-90-R).

### 3.2. Study Population

A total of eighty-two college students (36 non/low-drinkers and 46 BDs) from the UM (Braga, Portugal), aged between 18 and 23 years and matched for age and gender, will be recruited for the study. The sample size was determined based on statistical power analysis for two groups (Control and BDs), using the G*Power software (Erdfelder et al., 2009). Thus, employing an α level of 0.05, an effect size of 0.6 (considered a medium effect size), and a desired power of 0.80, the sample size for the control group was determined to be 36 subjects. Similarly, applying these parameters to the analysis of the BD group, a sample size of 19 for each BD subgroup (inulin intervention and maltodextrin intervention) was obtained. To account for an estimated dropout rate of approximately 4 individuals for each BD subgroup, the adjusted sample size was set to 23 subjects per group, resulting in a total of 46 BD subjects.

Participants who report (i) consuming 5 or more drinks on one occasion at least once a month, and (ii) drinking at a rate of at least two drinks per hour during these episodes, will be classified as BDs. Those who (i) never drank 5 or more drinks on any occasion, and (ii) had an AUDIT score ≤ 4, will be considered non/low-drinkers. The exclusion criteria will be as follows: use of illegal drugs except cannabis (i.e., at most once a month); alcohol abuse (i.e., score ≥20 in the AUDIT); consumption of medical drugs with psychoactive effects (e.g., sedatives or anxiolytics) during the two weeks before the experiment; personal history of psychopathological disorders (according to DSM-5 criteria); history of traumatic brain injury or neurological disorder; family history of substance abuse (including alcoholism); occurrence of an episode of loss of consciousness for more than 30 minutes; non-corrected sensory deficits; score ≥ 90 in the Global Severity Index of SCL-90-R or in at least two of the symptomatic dimensions; use of any of the following drugs in the last 4 weeks: laxatives, antibiotics, anticoagulants, non-steroidal anti-inflammatory drugs, analgesics, corticosteroids; as well as prebiotics and probiotics; diagnosis of any gut disease/problems or other medical conditions, such as inflammatory bowel disease, irritable bowel syndrome, Crohn’s Disease, celiac disease, lactose intolerance, and autoimmune disease.

### 3.3. Randomization and Blinding

The intervention protocol with prebiotic or placebo will be implemented by a qualified research assistant not otherwise involved in the study, who will pack the prebiotic and placebo capsules and randomly distribute them to each BD subgroup. For six weeks, BDs will receive a daily dose of 15 g divided into three equal administrations per day of one of the following interventions: (1) Prebiotic inulin intervention (n = 23) or (2) placebo intervention with maltodextrin (n = 23). The treatment groups will remain undisclosed to both researchers and participants until the final analyses.

## 4. ASSESSMENT PROTOCOL

### 4.1. Neuropsychological Evaluation

During both the pre-intervention (for all participants) and post-intervention (only for BDs) phases, participants will perform a number of neuropsychological tasks through a touch-sensitive screen (iPad) using the CANTAB, a battery of computerized tests that provide a comprehensive evaluation of cognitive functions which has been validated for clinical and research purposes (Boyle et al., 2023). The neurocognitive assessment will focus on executive functions (i.e., working memory, inhibitory control, cognitive flexibility, and decision-making), as well as short-term memory and emotion recognition abilities (Table 1).

### 4.2. Gut Microbiota

#### 4.2.1. Stool samples

Stool samples will be collected from the control (pre-intervention) and the BD (pre- and post-intervention) groups for gut microbiota diversity/composition analysis by 16S rRNA metagenomics (Illumina sequencing). Each participant’s sample will be collected with an EasySampler® Stool Collection Kit, kept in anaerobioses, and refrigerated until delivery to the laboratory. The sample will be aliquoted at the Centre of Biological Engineering (UM) and stored at – 80 °C for later analysis. Participants will also be instructed to collect the stool sample as close as possible to the pre- or post-intervention visit, with a maximum of 24 hours prior.

Stoll samples will be further divided in aliquots of 1 g, suspended in sterile 0.1 M phosphate-buffered saline (PBS) at 10% and stored at - 20 °C until the DNA extraction. Further, the genomic DNA will be extracted and purified from stool samples using the NZY Tissue gDNA Isolation Kit (Nzytech, Portugal), with some modifications (Roupar et al., 2022). DNA purity and quantification will be evaluated with a NanoDrop spectrophotometer. Further, sequencing of the bacterial 16s rRNA V3-V4 gene region will be performed by RTL Genomics (Lubbock, TX, USA) on the Illumina MiSeq platform (Illumina Inc., San Diego, CA, USA).

To understand the impact of prebiotic administration on the gut microbiota, all participants in the intervention group will be required to complete a food diary before and during the intervention. Wherein this diary, they will detail all the food and drink they consumed over three days of each intervention week (two weekdays and one weekend), including the type of packaging and the location where the food or beverage was consumed.

#### 4.2.2. Blood samples - Inflammatory markers

Blood samples will be collected by a qualified nurse at the EPsi (UM) during both pre- and post-intervention phases. Participants will be instructed to fast at the time of collection. Blood will be stored in BD Vacutainer® tubes containing Ethylenediamine Tetraacetic Acid (EDTA). Subsequently, the plasma will be aliquoted and preserved at – 80 °C for later analysis. Peripheral levels of the interleukins IL-1β, IL-6, and IL-8, and the TNF-α will be determined by enzyme-linked immunosorbent assay (ELISA) kits.

### 4.3. Neuroimaging assessment

In the pre- (for all participants) and post-intervention (only for BDs) phases, a neurostructural and neurofunctional assessment will be conducted at the Hospital da Luz (Guimarães, Portugal), by technical staff specialized in radiology and nursing, in collaboration with the research team. Participants will be instructed to abstain from consuming alcohol within the last 24 hours and to avoid BD episodes for the 3 days preceding the MRI session. Before scanning, potential alcohol use will be tested using a Breathalyzer device, and the assessment will only proceed after confirming 0% breath alcohol level. Furthermore, participants will be asked not to smoke or consume tea or coffee for at least 3 hours before the experiment.

Recordings will be conducted in a 3 T Magneton Trio (Siemens) clinical MRI scanner equipped with an 8-channel receive-only head coil. Participants will wear earmuffs to reduce the scanner noise and head motion will be minimized using foam pads. The neuroimaging recording protocol will include structural (T1-weighted and diffusion-weighted imaging [DWI]) and functional (T2*-weighted) acquisitions. The total acquisition time will be approximately 57 minutes, divided into 5.35 mins of T1, 14.12 mins DWI, 7.03 mins of resting-state fMRI, and 30.32 for tasks-related acquisitions. The total duration of the experiment (including preparation, acquisition protocol, and tasks performance) will be around 100 mins. During the tasks-related fMRI acquisitions, participants will view images of experimental tasks on a large monitor at the opening of the MRI scanner, using a mirror placed over their face. The monitor will display images in reverse so that they appeared normal when viewing through the mirror. In addition, participants will use a four-button fiber optic response pads (MRI compatible ResponseGrips, Nordic Neurolab) to record behavioural responses. Further details on MRI signal acquisition and analyses are described in Supplementary material.

#### 4.3.1. Experimental tasks (Figure 2, Image files document)

##### Think/No-Think Alcohol (TNTA) Task

The TNTA task will assess memory inhibition mechanisms in alcohol-related contexts (López-Caneda, Crego, et al., 2019). This task includes 18 images of beverages (9 alcoholic and 9 non-alcoholic) from the Galician Beverage Picture Set (López-Caneda & Carbia, 2018) and 18 images of human faces with a neutral expression (distributed equally by three age groups: young, middle-aged, and older), obtained from the Radboud Faces Database (Langner et al., 2010). The task includes 6 types of beverages: 3 alcoholic (beer, wine, and liquor) and 3 non-alcoholic (water, juice, and milk). Each image pair includes an image of a neutral human face and an image of a drink. The TNTA task comprises three phases: Learning, Think/No-Think (TNT), and Memory-Test. During the Learning phase, participants are asked to memorize 18 image pairs, divided into three blocks of 6 pairs each. Each block starts with a sequential random display of 6 image pairs on the screen for 4 s. Afterwards, only the human faces are shown, and participants must attempt to recall the alcoholic or non-alcoholic image that was associated with the neutral face by answering two questions: “Which drink was associated with this image?”; “How many people were in the picture?”. In each block, the 6 image pairs and the questions are repeated three times. At the end of each block, participants receive feedback on the number of correct answers. An answer is considered correct only if participants respond correctly to both questions. In the TNT phase, only the faces with neutral expressions are shown. Participants are instructed to either *think* (indicated by a green frame surroding the face)—meaning they should focus on the face and recall the associated alcoholic or non-alcoholic image -- or to *no-think* (indicated by a red frame surroding the face) —meaning they should focus on the face and actively prevent the previously associated picture from entering their consciousness. During this phase, only 12 faces are depicted, as the remaining 6 serve as a baseline condition for the following phase. Finally, in the Memory-Test phase, all 18 neutral faces from the initial pairs are presented again and participants are asked to remember the image (alcoholic or non-alcoholic) that was originally associated with each face, as indicated by the two questions from the Learning phase.

**Figure 2.**
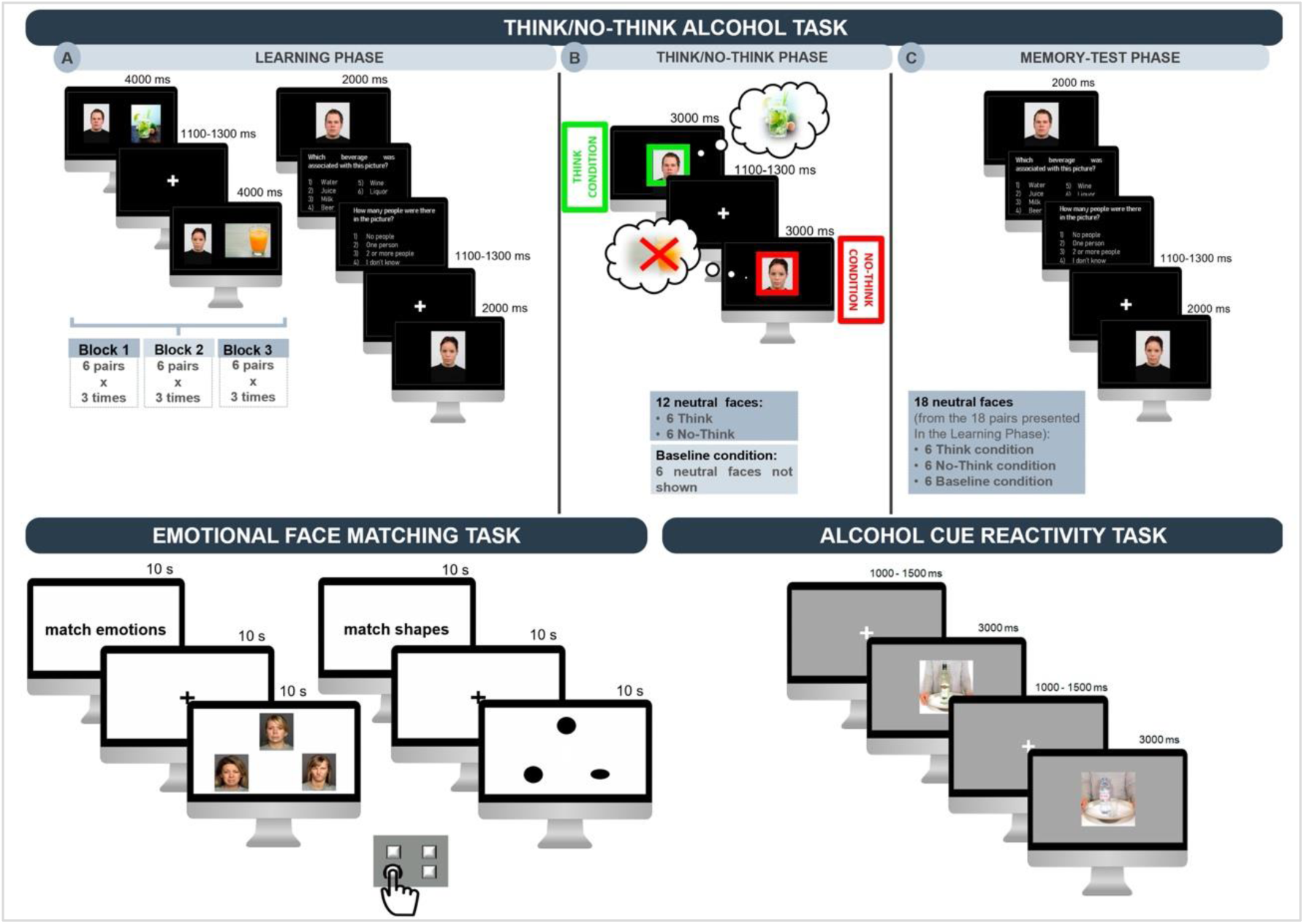
Graphic representation of the three tasks used during brain activity recordings in the MRI scanner: the Think/No-Think Alcohol task (top), the Emotional Face Matching task (bottom left), and the Alcohol Cue Reactivity task (bottom right).

##### Emotional Face Matching (EFM) task

The ability to recognize emotions will be assessed through the EFM task (Hariri et al., 2000). This paradigm assesses the brain’s preconscious and conscious responses to emotional faces, using faces with emotional expressions (anger, sadness, and fear) —obtained from the FACES database (Ebner et al., 2010)- for the experimental condition (match emotions) and geometric forms (ovals or circles) for the control condition (match shapes). Consisting of 3 face matching blocks alternated with 3 control blocks, without breaks in between, this task lasts 10 min. Each block comprises 18 trials, presented for 5 s with no interstimulus interval. Instructions (“match emotions” or “match shapes”) are displayed for 10 s, followed by a 10-second crosshair fixation before the next block begins. During each trial, participants view three emotional faces or shapes (one on top and two on the bottom, in a triangular configuration) and are instructed to identify the emotion/shape at the top of the screen and match it with the corresponding one at the bottom using handheld response buttons.

##### Alcohol Cue Reactivity (ACR) task

The reactivity to alcohol cues will be assessed using the ACR task (Cofresí et al., 2024). This task starts with a white fixation cross displayed on a grey background for a variable duration ranging from 100 to 150 ms. Subsequently, either an alcoholic or non-alcoholic drink is randomly presented at the centre of the screen for 300 ms. Participants are instructed to attentively observe all images presented on the screen. Similar to the study by (Wiers et al., 2015), designed to assess participants’ task engagement, four oddball blocks were included. Each oddball block consists of four alcoholic or non-alcoholic stimuli interspersed with an oddball cue—a picture of a neutral object (e.g., a button or a padlock). During these blocks, participants are required to press a button using their right hand. After viewing all images, participants are asked to rate their emotional responses to the alcoholic or non-alcoholic beverage images in terms of valence, arousal, and desire, using the Self-Assessment Manikin (Bradley & Lang, 1994). The complete task encompasses a total of 80 beverage images (40 alcoholic and 40 non-alcoholic), sourced from the Amsterdam Beverage Picture Set (Pronk et al., 2015) and neutral objects images from the POPORO database (Kovalenko et al., 2012).

## 5. STATISTICAL ANALYSIS PLAN

Regarding statistical analysis concerning GM, the primary outcome will assess between-group differences at two levels: α- (within-sample) and β- (between-sample) diversity (Kers & Saccenti, 2022). Measures of *α*-diversity will include: 1) total number of differential abundance of microbial species; 2) Chao1 index (a nonparametric richness estimator); 3) Shannon diversity index (a metric combining richness and evenness, assigning equal weight to abundant and rare species); and 4) Simpson diversity index (a metric of richness and evenness, giving more weight to abundant species). Measures of β-diversity -i.e., the similarity or distance between microbiome pairs-will include Bray– Curtis dissimilarity (a measure of community composition differences), and weighted and unweighted UniFrac (indexes of phylogenetic community differences).

Data on the number of copies achieved will be normalized by the mean number of copies of 16S rRNA for each microorganism detected. Microbial community analysis will be performed by sequencing the 16S rRNA gene (V4 region) using the prokaryotic universal primer pair 515f/806r (Caporaso et al., 2012), by MiSeq Illumina sequencing at Research and Testing Laboratory (Lubbock, TX), as described by (Salvador et al., 2018). Detailed information on the bioinformatics analysis steps can be found on the RTL website (https://rtlgenomics.com/amplicon-bioinformatics-pipeline). Microbiota composition (highlighting the dominant phyla: Firmicutes, Bacteroides, Actinobacteria, and Proteobacteria) will be correlated with data obtained at the beginning and end of the intervention. According to open science policy, sequencing data (FASTQ files) from the microbiome will be deposited in public databases such as the European Nucleotide Archive of the European Bioinformatics Institute (ENA-EBI).

The study also aims to characterize the presence of some immune/inflammatory markers (the TNF-α and the interleukins IL-1β, IL-6, and IL-8) in the blood of individuals with BD in comparison with the control group. First, the data will be evaluated for normality using the Shapiro-Wilk test. If the data are normal, the Student t-test for independent samples will be used to compare the levels of immune/inflammatory markers between non/low drinkers and BDs or between the inulin and maltodextrin intervention group. Otherwise, for data without normal distribution, the Mann-Whitney test will be used. To investigate correlations between levels of inflammatory biomarkers and other relevant variables, we will use the Pearson correlation coefficient for data with normal distribution and the Spearman coefficient for non-parametric data. A significance level of 0.05 will be adopted for all analyses and comparisons.

Neuropsychological performance will be assessed through the Delayed Matching to Sample, Emotion Recognition, Cambridge Gambling, Intra-Extra Dimensional Set Shift, Spatial Working Memory, and Stop Signal tasks. Following the same parameters as the analysis of pro-inflammatory markers, we will conduct independent sample t-tests to explore potential mean differences between BDs and non/low drinkers on key outcome measures previously identified for each task (see CANTAB Connect Research Outcome Measures in Supplementary Material), including the number of correct responses, mean reaction times, commission/omission errors, etc.

For structural and resting-state fMRI data, multivariate analysis of variance and two-way mixed ANOVAs will be applied with group (non/low drinkers vs. BDs) and gender (male vs. female) as the between-subjects factors, hemisphere (left vs. right) as the within-subjects factor, and age as covariate to test for differences in grey and white matter volumes, functional connectivity, and resting-state maps. For these and the remaining analyses, degrees of freedom will be corrected by the Greenhouse-Geisser procedure when appropriate. Family-wise error (FWE) corrections will be applied to both main and interaction effects and post-hoc paired comparisons will be conducted with the Bonferroni adjustment for multiple comparisons (α ≤ 0.05). Regarding task-related brain activity, the contrast images from the first-level analysis will be used for the second-level analysis. We will analyse the data using a repeated measures analysis of variance (ANOVA) for each task to assess the main effects of group (i.e., non/low drinkers vs. BDs), conditions (e.g., alcohol vs. non-alcohol, think vs. no-think), and the group x conditions interactions using the flexible factorial approach in SPM12. A whole-brain cluster family-wise error (FWE) correction at p < .05 will be applied, with an uncorrected voxel-level cluster-defining threshold of p < .001.

To explore the complex relationships among alcohol use, microbial diversity/composition, neuropsychological measures, proinflammatory markers, and structural and functional brain characteristics, we will develop a Structural Equation Modeling (SEM) (Ullman, 2006). This model will encompass six primary latent variables: Alcohol Use Patterns, Gut Microbiota, Neuropsychological Performance, Immune Response, Brain Structure, and Brain Functioning. Each of these latent variables will be operationalized by the aforementioned observed variables (e.g., membership in the BD or non/low drinkers group, α-/β-diversity, levels of TNF-α and interleukins, volumes of gray and white matter). The proposed SEM model will investigate regression relationships among these latent variables to ascertain how gut microbiota, through its diversity and composition, could indirectly influence brain and behavior. Additionally, covariances between latent variables will be examined to elucidate interactions among the various dimensions of the study.

Finally, the RCT will aim to determine whether prebiotic administration in a subgroup of BDs can affect the microbiota diversity/composition and if these changes may influence other health measures such as inflammatory response or brain volume. To this end, we will employ a mixed-effects linear regression model, aimed at assessing how treatment (prebiotic vs. placebo) affects the outcomes related to immune response, neuropsychological performance, and brain structure/functioning across a sample of individuals characterized by BD behavior.

## 6. DISCUSSION

To the best of our knowledge, this protocol will be the first to explore the potential relationship between the *Brain* (neurocognitive functioning), the *Bugs* (gut microbiota), and the *Binge* (alcohol use) in a population of youths with a BD pattern in comparison with youths with low or no alcohol use. This study will also implement a RCT aimed at exploring the potential effects of a prebiotic intervention along the microbiota-gut-brain axis in this population. In addition to representing an innovative step, this approach provides an opportunity to broaden the understanding of changes in the gut microbiome in BDs, as well as their behaviour and neurocognitive functioning.

The uniqueness of this protocol lies in the exploration of the complex interaction between alcohol, brain activity, and gut microbiome through the implementation of multiple levels of analysis, including techniques to measure brain activity (i.e., MRI), paradigms to measure cognitive performance, stool, and blood sampling, and the application of various questionnaires concerning psychological and behavioural characteristics. By integrating these diverse methodologies, we aim to elucidate the intricate mechanisms underlying the effects of alcohol consumption on both the brain and the gut microbiome, shedding light on potential biomarkers and cognitive changes associated with different levels of alcohol intake. This comprehensive approach not only allows for a more holistic understanding of alcohol’s impact on human physiology and cognition but also opens up new avenues for tailored interventions and preventive strategies aimed at mitigating the negative consequences of excessive alcohol consumption.

## 7. ETHICS AND DISSEMINATION

The Ethics Committee for Social and Human Sciences of University of Minho approved the present protocol on June 28, 2022 (approval reference: CEICSH 078/2022), ensuring that all aspects of research will be followed in accordance with the national and international accepted guidelines and standards – of the Code of Ethical Principles for Medical Research Involving Humans Subjects outlined in the Declaration of Helsinki (64th World Medical Association General Assembly, Brazil, 2013).

The data will be treated as confidential and will solely be used for research purposes by the research team, complying with General Data Protection Regulations (GDPR). The research team will underscore that participation is voluntary, they can withdraw from the study at any time. Before enrolling in the study, all participants will be informed about the aims, conditions, and procedure of the study and provided with two copies of the informed consent forms signed by the researchers and participants. College students will receive gift vouchers to compensate for their participation.

Ethical issues identified are related to the collection and processing of sensitive personal health/lifestyle data from participants. The project uses safe and non-invasive methods. The prebiotics used in this randomized clinical trial are proven to have no side/negative effects (Sheng et al., 2023), as they are classified as food ingredients in Europe (Victoria Obayomi et al., 2024), hold GRAS (Generally Recognized as Safe) status in the United States (Burdock & Carabin, 2004), and are recognized as dietary fibers by the Food and Drug Administration (Slavin, 2013).

Upon thorough analysis of the data from this project, regardless of the outcome, the findings will be presented at national and international conferences and published in peer-reviewed scientific journals indexed in the Journal Citation Reports, preferentially in open-access formats. Additionally, data may be shared with third parties solely for research purposes and upon reasonable request, in line with ethical guidelines and data protection regulations. The anonymity of all participants will be strictly maintained, and all data will be handled in compliance with relevant data protection laws, including the GDPR.

## Supporting information

SPIRIT 2013 Checklist;MRI acquisition;MRI processing and analysis; CANTAB Measures; Contingency Plan or Risk Analysis; Informed Consent Form

## Data Availability

All data produced in the present work are contained in the manuscript.
URLs/accession numbers/DOIs will be available only after acceptance of the manuscript for publication in BMJ Journals, so that we can ensure their inclusion before publication.

## ACKNOWLEDGEMENTS

This study is being conducted at the Psychology Research Center, School of Psychology, University of Minho supported by the Portuguese Foundation for Science and Technology (FCT) through the Portuguese State Budget (UIDB/PSI/01662/2020), in collaboration with the Centre of Biological Engineering (UIDB/04469/2020). This study is also supported by the project PTDC/PSI-ESP/1243/2021, funded by the FCT. Eduardo López Caneda and Clarisse Nobre acknowledge the FCT for the Assistant Research contracts CEECIND/07751/2022 and 10.54499/2021.01234.CEECIND/CP1664/CT0019, respectively. Alberto Crego is supported by the FCT and the Portuguese Ministry of Science, Technology and Higher Education, within the scope of the Transitory Disposition of the Decree No. 57/2016. Carina Carbia benefited from funding by the Fonds de la Recherche Scientifique – FNRS, Chargé de Recherches (CR). Natália Almeida-Antunes was supported by the FCT, MCTES, and the European Union through the European Social Fund (SFRH/BD/146194/2019).

