## Supplementary material for "The Brain, the Bugs, and the Binge: A Protocol Intervention Examining the Interplay Between Binge drinking, Gut Microbiota, and Brain Functioning": SPIRIT 2013 Checklist;MRI acquisition;MRI processing and analysis; CANTAB Measures; Contingency Plan or Risk Analysis; Informed Consent Form: Supplementary Material.pdf

### Standard Protocol Items for Randomized Trials (SPIRIT) 2013 Checklist

| Section/item | Item No | Ms Pg | Description |
|---|---|---|---|
| **Administrative information** | | | |
| Title | 1 | 1 | Descriptive title identifying the study design, population, interventions, and, if applicable, trial acronym |
| Trial registration | 2a | 3 | Trial identifier and registry name. If not yet registered, name of intended registry |
| | 2b | 1-30 | All items from the World Health Organization Trial Registration Data Set |
| Protocol version | 3 | 8-12 | Date and version identifier |
| Funding | 4 | 21-22 | Sources and types of financial, material, and other support |
| Roles and responsibilities | 5a | 1 | Names, affiliations, and roles of protocol contributors |
| | 5b | 1 | Name and contact information for the trial sponsor |
| | 5c | 1 | Role of study sponsor and funders, if any, in study design; collection, management, analysis, and interpretation of data; writing of the report; and the decision to submit the report for publication, including whether they will have ultimate authority over any of these activities |
| | 5d | 1 | Composition, roles, and responsibilities of the coordinating centre, steering committee, endpoint adjudication committee, data management team, and other individuals or groups overseeing the trial, if applicable (see Item 21a for data monitoring committee) |
| **Introduction** | | | |
| Background and rationale | 6a | 4-8 | Description of research question and justification for undertaking the trial, including summary of relevant studies (published and unpublished) examining benefits and harms for each intervention |
| | 6b | 4-7 | Explanation for choice of comparators |
| Objectives | 7 | 7-8 | Specific objectives or hypotheses |
| Trial design | 8 | 2-3 | Description of trial design including type of trial (eg, parallel group, crossover, factorial, single group), allocation ratio, and framework (eg, superiority, equivalence, noninferiority, exploratory) |
| **Methods: Participants, interventions, and outcomes** | | | |
| Study setting | 9 | 8-11 | Description of study settings (eg, community clinic, academic hospital) and list of countries where data will be collected. Reference to where list of study sites can be obtained |
| Eligibility criteria | 10 | 11-12 | Inclusion and exclusion criteria for participants. If applicable, eligibility criteria for study centres and individuals who will perform the interventions (eg, surgeons, psychotherapists) |
| Interventions | 11a | 9-11 | Interventions for each group with sufficient detail to allow replication, including how and when they will be administered |
| | 11b | 11-12 | Criteria for discontinuing or modifying allocated interventions for a given trial participant (eg, drug dose change in response to harms, participant request, or improving/worsening disease) |
| | 11c | A4 | Strategies to improve adherence to intervention protocols, and any procedures for monitoring adherence (eg, drug tablet return, laboratory tests) |

| | | | |
|---|---|---|---|
| | 11d | 10 | Relevant concomitant care and interventions that are permitted or prohibited during the trial |
| Outcomes | 12 | 17-20 | Primary, secondary, and other outcomes, including the specific measurement variable (eg, systolic blood pressure), analysis metric (eg, change from baseline, final value, time to event), method of aggregation (eg, median, proportion), and time point for each outcome. Explanation of the clinical relevance of chosen efficacy and harm outcomes is strongly recommended |
| Participant timeline | 13 | 8-11 | Time schedule of enrolment, interventions (including any run-ins and washouts), assessments, and visits for participants. A schematic diagram is highly recommended (see Figure) |
| Sample size | 14 | 11-12 | Estimated number of participants needed to achieve study objectives and how it was determined, including clinical and statistical assumptions supporting any sample size calculations |
| Recruitment | 15 | 9 | Strategies for achieving adequate participant enrolment to reach target sample size |
| **Methods: Assignment of interventions (for controlled trials)** | | | |
| Allocation: | | | |
| Sequence generation | 16a | 11-12 | Method of generating the allocation sequence (eg, computer-generated random numbers), and list of any factors for stratification. To reduce predictability of a random sequence, details of any planned restriction (eg, blocking) should be provided in a separate document that is unavailable to those who enrol participants or assign interventions |
| Allocation concealment mechanism | 16b | 11-12 | Mechanism of implementing the allocation sequence (eg, central telephone; sequentially numbered, opaque, sealed envelopes), describing any steps to conceal the sequence until interventions are assigned |
| Implementation | 16c | 12 | Who will generate the allocation sequence, who will enrol participants, and who will assign participants to interventions |
| Blinding (masking) | 17a | 12 | Who will be blinded after assignment to interventions (eg, trial participants, care providers, outcome assessors, data analysts), and how |
| | 17b | 12 | If blinded, circumstances under which unblinding is permissible, and procedure for revealing a participant's allocated intervention during the trial |
| **Methods: Data collection, management, and analysis** | | | |
| Data collection methods | 18a | 12-17, A1, A2, A3 | Plans for assessment and collection of outcome, baseline, and other trial data, including any related processes to promote data quality (eg, duplicate measurements, training of assessors) and a description of study instruments (eg, questionnaires, laboratory tests) along with their reliability and validity, if known. Reference to where data collection forms can be found, if not in the protocol |
| | 18b | A4 | Plans to promote participant retention and complete follow-up, including list of any outcome data to be collected for participants who discontinue or deviate from intervention protocols |
| Data management | 19 | 20-21 | Plans for data entry, coding, security, and storage, including any related processes to promote data quality (eg, double data entry; range checks for data values). Reference to where details of data management procedures can be found, if not in the protocol |
| Statistical methods | 20a | 17-19 | Statistical methods for analysing primary and secondary outcomes. Reference to where other details of the statistical analysis plan can be found, if not in the protocol |
| | 20b | 17-19 | Methods for any additional analyses (eg, subgroup and adjusted analyses) |

| | | | |
|---|---|---|---|
| | 20c | | Definition of analysis population relating to protocol non-adherence (eg, as randomised analysis), and any statistical methods to handle missing data (eg, multiple imputation) |
| **Methods: Monitoring** | | | |
| Data monitoring | 21a | 1 | Composition of data monitoring committee (DMC); summary of its role and reporting structure; statement of whether it is independent from the sponsor and competing interests; and reference to where further details about its charter can be found, if not in the protocol. Alternatively, an explanation of why a DMC is not needed |
| | 21b | | Description of any interim analyses and stopping guidelines, including who will have access to these interim results and make the final decision to terminate the trial |
| Harms | 22 | A4 | Plans for collecting, assessing, reporting, and managing solicited and spontaneously reported adverse events and other unintended effects of trial interventions or trial conduct |
| Auditing | 23 | A4 | Frequency and procedures for auditing trial conduct, if any, and whether the process will be independent from investigators and the sponsor |
| **Ethics and dissemination** | | | |
| Research ethics approval | 24 | 20-21 | Plans for seeking research ethics committee/institutional review board (REC/IRB) approval |
| Protocol amendments | 25 | A4 | Plans for communicating important protocol modifications (eg, changes to eligibility criteria, outcomes, analyses) to relevant parties (eg, investigators, REC/IRBs, trial participants, trial registries, journals, regulators) |
| Consent or assent | 26a | 10-11 | Who will obtain informed consent or assent from potential trial participants or authorised surrogates, and how (see Item 32) |
| | 26b | | Additional consent provisions for collection and use of participant data and biological specimens in ancillary studies, if applicable |
| Confidentiality | 27 | 20-21 | How personal information about potential and enrolled participants will be collected, shared, and maintained in order to protect confidentiality before, during, and after the trial |
| Declaration of interests | 28 | 22 | Financial and other competing interests for principal investigators for the overall trial and each study site |
| Access to data | 29 | | Statement of who will have access to the final trial dataset, and disclosure of contractual agreements that limit such access for investigators |
| Ancillary and post-trial care | 30 | | Provisions, if any, for ancillary and post-trial care, and for compensation to those who suffer harm from trial participation |
| Dissemination policy | 31a | 20-21 | Plans for investigators and sponsor to communicate trial results to participants, healthcare professionals, the public, and other relevant groups (eg, via publication, reporting in results databases, or other data sharing arrangements), including any publication restrictions |
| | 31b | | Authorship eligibility guidelines and any intended use of professional writers |
| | 31c | | Plans, if any, for granting public access to the full protocol, participant-level dataset, and statistical code |
| **Appendices** | | | |
| Informed consent materials | 32 | A5 | Model consent form and other related documentation given to participants and authorised surrogates |

| | | | |
|---|---|---|---|
| Biological specimens | 33 | 13-14 | Plans for collection, laboratory evaluation, and storage of biological specimens for genetic or molecular analysis in the current trial and for future use in ancillary studies, if applicable |

*It is strongly recommended that this checklist be read in conjunction with the SPIRIT 2013 Explanation & Elaboration for important clarification on the items. Amendments to the protocol should be tracked and dated. The SPIRIT checklist is copyrighted by the SPIRIT Group under the Creative Commons "Attribution-NonCommercial-NoDerivs 3.0 Unported" license.

### A1. Appendix 1: MRI acquisition

Sagittal high-resolution 3D T1 weighted anatomical images will be acquired using a magnetization prepared rapid acquisition gradient echo (MPRAGE) sequence with the following parameters: repetition time (TR) = 2400 ms, echo time (TE) = 2.53 ms, inversion time (TI) = 1.100 ms, flip angle (FA) = 9°, 176 slices with 0.98 mm thickness, slab thickness = 172.48 mm, slice gap = 0 mm, in-plane resolution = 1x1 $mm^2$, matrix size = 256x256 and 250mm FoV.

The Diffusion Weighted Imaging (DWI) will be performed using a spin-echo echo-planar imaging (SE-EPI) sequence: TR = 6200 ms; TE = 96 ms; FoV read=220 mm; acquisition matrix = 91×110; 44 slices, slice thickness = 3.5 mm; 64 non-collinear gradient directions with b = 1000 s/mm2; and two b = 0 s/mm2 acquisition, slice gap = 0 mm, voxel size = 2.4x2x3.5 mm.

The blood oxygen level-dependent (BOLD) sensitive echo-planar imaging (EPI) will be used with the following acquisition parameters: 48 interleaved axial slices; TR = 2000ms; TE = 18ms; FA = 85º; slice thickness = 3mm; voxel size = 4.1x3x3mm; FoV = 200mm. matrix size: 49x66, slice gap = 0 mm.

The resting state fMRI will be performed using a blood oxygen level dependent (BOLD) sensitive echo-planar imaging (EPI) with the parameters: 46 interleaved axial slices, TR = 1460 ms, TE = 12 ms, flip angle (FA) = 85°, slice thickness = 3 mm, voxel size = 5.1x3x3 mm, FoV = 200 mm and 285 volumes.

### A2. Appendix 2: MRI processing and analysis

After the acquisition, all MRI scans will be visually inspected to discard critical head motion or brain lesions. After this verification, each subject's T1-weighted MRI will be processed using the automated reconall FreeSurfer preprocessing, parcellation, and segmentation standard steps (http://surfer.nmr.mgh.harvard.edu) to obtain the cortical surface reconstruction and tissue-class segmentation. Briefly, the pipeline will include motion correction and intensity normalization, removal of non-brain tissue and skull stripping, Talairach transformations, whole-brain segmentation, cortical parcellation, and statistical outputs.

DTI preprocessing will be performed with tools provided by the FMRIB Software Library (FSL- http://fsl. fmrib.ox.ac.uk/fsl/). The analysis pipeline will be carried out as follows: i) motion and eddy current corrections using FMRIB's Diffusion Toolbox (FDT); ii) brain extraction tool (BET) applied to the T1 and DWI images; iii) tensor fitting computation will be performed with DTIFIT; iv) after tensor estimation, tractography and scalar maps of FA, AD, RD and MD will be generated.

Functional imaging data preprocessing and analysis will be performed with Statistical Parametric Mapping SPM12 (Wellcome Trust Centre for Neuroimaging, London, UK). In the preprocessing, all functional images will be corrected for slice timing and motions using realign, co-registered to T1-weight structural image and spatially normalized into the Montreal Neurological Institute (MNI) template. These normalized images will be spatially smoothed using a Gaussian kernel of 8-mm full width at half maximum (FWHM) and low-frequency drifts will be removed with a high-pass temporal filter (filter width of 128s).

### A3. Appendix 3: CANTAB Connect Research Outcome Measures

Six tasks of the CANTAB will be administered to evaluate different components of cognitive functions, and task-specific performance measures related to speed as well as task performance accuracy will be analysed to provide a comprehensive profile of cognitive function. Table 1 describes the outcome measures used in the analysis of each task.

**Table 1.** Outcome measures descriptions of the cognitive tasks from the Cambridge Neuropsychological Tests Automated Battery (CANTAB).

| **Measure Code** | **Description** |
|---|---|
| *Delayed Matching to Sample* | |
| DMSPCAD<br>*Percent Correct All Delays* | Percentage of success in trials in which the subject chose the correct pattern on the first attempt. Calculated for all trials. |
| DMSPCS, DMSPCO, DMSPC4, DMSPC12<br>*Percent Correct Simultaneous/0/4/12* | Percentage of success calculated in all trials in which there was an interval of zero, four and twelve seconds between stimulus and response hypotheses. |
| *Emotion Recognition Task* | |
| ERTCRT H/S/F/A/D/SU<br>*Median Correct Reaction Time* | Reaction time of the correct choice of emotion after being presented with a face, calculated in all tests evaluated. |
| ERTUHR H/S/F/A/D/SU<br>*Unbiased Hit Rate* | Unbiased hit rate, which ensures that the accuracy of emotion recognition is not influenced by response guessing or response bias effects. |
| ERTTHH/S/F/A/D/SU<br>*Total Hits* | The number of times the subject correctly selected the emotion in all trials evaluated. |
| ERTTFAH S/F/A/D/SU<br>*Total False Alarms* | The number of times the subject has incorrectly selected an emotion (false alarms). |
| *Cambridge Gambling Task* | |
| CGTDMQMT<br>*Decision Making Quality Total Merged* | The ratio (0 – 1) of trials in which the subject chose the colour of the majority squares, calculated for all trials (up and down) in which the number of squares of each colour differed. |
| CGTRAJTM<br>*Risk Adjustment Merged* | Measure of risk adjustment calculated from the average proportion of points that the subject has chosen to bet, taking into account the majority of coloured squares. |
| CGTDAVT<br>*Delay Aversion Total* | A measure that allows the dissociation between risk taking and impulsiveness, determining whether the subject made a bet at the first opportunity. Calculated based on the subtraction of risk taking in all ascending trials to risk taking in descending trials. |
| *Intra-Extra Dimensional Set Shift* | |
| IEDEEDS<br>*EDS Errors* | The number of times that the subject failed to select the rule-compatible stimulus in the phase in which the extradimensional displacement occurs. A measure of the subject’s ability to change the focus of attention. |

| | |
|---|---|
| IEDYERTA<br>*Total Errors Adjusted* | The number of times the subject chose a wrong stimulus (incompatible with the current rule, adjusted for each stage of the task that was not reached). |
| *Spatial Working Memory* | |
| SWMS<br>*Strategy* | The number of times the subject starts new research. It provides information about strategy. |
| SWMBE468<br>*Between Errors* | The number of times the subject incorrectly revisits a square in which a token was previously found. Calculated on all trials evaluated (with four, six, and eight tokens). |
| SWMBE4 /SWMBE6 /SWMBE8<br>*Between Errors 4/6/8 Boxes* | The number of times the subject revisits a square in which a token was previously found. Calculated for each type of test (4, 6, or 8 tokens). |
| *Stop Signal Task* | |
| SSTSSRT<br>*Reaction Time* | Reaction time for trials in which the participant successfully inhibited the response. |

**Note**: H= Happiness; F= Fear; S= Sadness; A = Anger; D = Disgust; SU = Surprise; DMS = Delayed Matching to Sample; SWM = Spatial Working Memory; ERT = Emotional Recognition Task; IED = Intra-Extra Dimensional Set Shift; SST = Stop Signal Task; CGT = Cambridge Gambling Task.

### A4. Appendix 4: Contingency Plan or Risk Analysis

1. **Recruitment of Participants**
   ***a) Selection Bias***
   *Risk:* Potential to select participants who do not adequately represent the target population, which could distort the results.
   *Solution:* Define clear and objective inclusion and exclusion criteria. Use diversified recruitment methods to reach a representative sample of the target population.

   ***b) Recruitment Difficulty***
   *Risk:* Possibility of not recruiting enough participants within the established timeframe, impacting study validity and completion.
   *Solution:* Conduct a preliminary assessment of participant availability and implement effective recruitment strategies, such as advertising to engage the entire university community.

2. **Logistical Difficulties in Sample Collection**
   *Risk:* Issues such as inadequate scheduling, lack of transportation to collection sites, long waits, or participant discomfort.
   *Solution:* Set flexible schedules for sample collection and imaging, provide transportation as needed, minimize waiting times, and ensure staff are available and responsive to participant needs and concerns.

3. **Biological Samples (Blood and Feces)**
   *Risk:* Potential sample contamination during collection, transport, or storage, compromising study results.
   *Solution:* Implement rigorous collection and handling procedures, including using sterile equipment and practicing all necessary techniques. Ensure proper storage conditions (e.g., controlled temperature).

4. **Participant Withdrawal**
   *Risk:* Participant withdrawal may lead to data loss and reduced statistical power.
   *Solution:* Use strategies to minimize dropouts, such as maintaining regular communication, providing incentives (e.g., vouchers), and ensuring a positive participant experience.

5. **Interruption of Prebiotic/Placebo Intervention**
   *Risk:* Mild side effects, forgetfulness, or non-adherence during the intervention phase.
   *Solution:* Closely monitor participants for any side effects and provide medical support as needed. Maintain regular contact to remind participants of treatment importance, offer incentives, and provide alternative delivery options (e.g., capsules or powder) to improve convenience and adherence.

### A5. Appendix 5: Informed Consent Form

**Official Title of the Study**:
The Brain, the Bug, and the Binge: a double-blind, randomized controlled trial investigating the interplay between binge drinking, gut microbiota and brain functioning.
**NCT ID**: NCT05946083
**Date**: October 28, 2024

University of Minho
School of Psychology
Research Center in Psychology (CIPsi)
Psychological Neuroscience Laboratory
Campus de Gualtar
4710-057 Braga

**Research Project reference**: PTDC/PSI-ESP/1243/2021
**Principal Investigator**: Dr. Eduardo López Caneda

#### Informed Consent Form

1) I confirm that I have read and understood the informative document that was delivered to me, containing all the information regarding the study in which I am participating, and that I have had the opportunity to raise questions and concerns about it.
2) I confirm that the research team has provided me with clear answers to all my questions and concerns.
3) I understand that I am free to leave the study at any time, without providing justification and without any consequences.
4) I understand and agree that my personal identification data and the data obtained through the course of the research study will be stored in separate archives to ensure its security, and that team members with access to the data will respect its confidentiality.
5) I, therefore, consent to my data being stored and/or exported to external databases for analysis, understanding that, under no circumstances, will information regarding my identity be disclosed.
6) I understand that this study does not have a diagnostic purpose and, consequently, I will not receive an individual report of my data/results.
7) I consent to participate in the above-mentioned study.

**Participant's name**: ..........................................................................................................

**Researcher's name:** ..........................................................................................................

**Name of Person Responsible for Collecting Consent** (if different from the Researcher)**:**

..........................................................................................................................................

**Date**: ..............................                **Participant's Code**: ......................................
